# Additive interaction and mediation-interaction decomposition: DNA methylation age acceleration, education, and cognitive impairment in the Health and Retirement Study

**DOI:** 10.1101/2022.12.23.22283919

**Authors:** Erin B. Ware, Cesar Higgins, Sean Harris, Jonah D. Fisher, Kelly M. Bakulski

## Abstract

**Importance:** Dementia represents a significant and increasing public health burden. DNA methylation age acceleration may be associated with dementia and dementia risk factors, such as education, but investigating their impact on dementia is necessary.

**Objective:** To evaluate the association of educational attainment on dementia and cognitive impairment through DNA methylation age acceleration, while accommodating exposure-mediator interaction effects.

**Design:** In the 2016 Health and Retirement Study wave, we evaluated six epigenetic clocks, including GrimAge, with Langa-Weir classified dementia, cognitive impairment non-dementia, and normal cognition. Age acceleration was the residual between chronological age of participants and DNA methylation clock, dichotomized at zero. To understand the joint association of low education (≤12 years) and DNA methylation age acceleration in relation to cognitive impairment, we used weighted logistic regression and calculated interaction on the additive scale adjusting for chronological age, sex, race/ethnicity, and cell type composition. We performed four-way mediation and interaction decomposition analysis to estimate the: 1) controlled direct effect of education on cognition, 2) interaction reference, 3) interaction mediation, and 4) the pure indirect effect of DNA methylation age acceleration on cognition.

**Setting:** Analysis was conducted on a subsample of Health and Retirement Study participants in the 2016 Venous Blood Study (N=3,724).

**Results:** Both GrimAge acceleration (OR=1.6 95%CI 1.3 – 2.1) and low educational attainment (OR=2.4 95%CI 1.9 – 3.0) were associated with higher odds of cognitive impairment, non-dementia in a mutually adjusted logistic model. We found additive interaction associations between low education and GrimAge acceleration on dementia. We observed that 6-8% of the association of education on dementia was mediated through GrimAge acceleration. While mediation effects were small, the portion of the association of education due to additive interaction with GrimAge acceleration was between 23.6 and 29.2%.

**Conclusions and Relevance:** Accelerated DNA methylation age was associated with increased odds of cognitive impairment and we observed more than additive interaction effects between education and age acceleration on dementia. These results support the interplay of social disadvantage and biological aging processes on impaired cognition.

---

Alzheimer’s disease and its related dementias are neurodegenerative diseases where the loss of cognitive function severely affects one’s daily activities. Dementia affects 5.7 million Americans, with rising global prevalence due to an aging population structure^1^. Currently, dementias are incurable, and annual care is estimated at $236 billion^2^. Disparities in dementia are linked to structural and socioeconomic factors including racism and level of education^3-7^. Novel approaches characterizing the multifaceted etiology of dementia are needed to identify the biological underpinnings and potentially modifiable environmental factors that lead to dementia disparities.

Epigenetics—modifications to the genome that are not changes in DNA sequence—are potential indicators of adverse changes at a molecular level, uncovering risk of disease before the outcomes are observable. Epigenetic measures, such as DNA methylation, measured in brain tissue are associated with dementia^8-10^. DNA methylation patterns in peripheral tissues, such as blood, are also associated with dementia risk factors and dementia outcomes^11-13^, providing a less invasive source for biomarkers compared to sources such as cerebrospinal fluid. These studies represent an important first step; however, foundational epigenetic epidemiology was primarily conducted in predominantly highly educated and white samples and clinical dementia case-control samples. It is crucial that we now examine larger, more socioeconomically and racially diverse samples.

Epigenetic aging clocks are molecular biomarkers of aging based on patterns of DNA methylation at cytosine-phosphate-guanosine sites in the genome^14^. Epigenetic clocks can be applied to DNA methylation obtained from whole blood. In fully understanding the efficacy of epigenetic clocks as a biomarker of dementia, it is critical to evaluate epigenetic clocks compared to chronological age and to evaluate how socioeconomic factors like educational attainment modify observed associations between epigenetic aging and dementia. The purpose of this study was to evaluate six different epigenetic clocks as predictors of dementia and cognitive impairment compared to chronological age and to investigate potential interaction with education. Additionally, we explored whether the epigenetic age acceleration mediates the association of education on cognitive impairment, while our mediation model accommodates for interaction effects between education and our epigenetic biomarker of interest. We used the Health and Retirement Study, a nationally representative study of adults over the age of 50 in the United States, to investigate the relationship between epigenetic age acceleration (through epigenetic clocks) and impaired cognition among individuals with varying education status (≤12 years of education, >12 years).

## METHODS

### Study population

The Health and Retirement Study is a nationally representative study sponsored by the National Institute on Aging (NIA U01AG009740). Surveys of adults over age 50 and their spouses in the United States are conducted by the University of Michigan, providing a national resource for data on changing health and economic circumstances associated with an aging population^15^. The Health and Retirement Study uses post-stratifying sampling weights methodology to obtain a representative sample of multiple birth cohorts of adults from two sexes and participants with diverse racial/ethnic backgrounds. Participants in the Health and Retirement Study provided written informed consent at the time of participation. Data collection procedures were approved by the University of Michigan Institutional Review Board. This secondary data analysis was approved by the University of Michigan Institutional Review Board (HUM00128220). All data used are publicly available through the Health and Retirement Study (https://hrs.isr.umich.edu/).

### DNA methylation assays

DNA methylation assays were conducted on a non-random subsample (n=4,104) of respondents who participated in the 2016 Venous Blood Study. This subsample fully represents the entire Health and Retirement Study sample. Venous blood was collected by Hooper Holmes Health and Wellness. The quantity of blood collected was 50.5 mL split into 6 tubes: 1 × 8 mL CPT tube, 3 × 10 mL double gel serum separator tubes (SST), 1 × 10 mL EDTA whole blood tube, and 1 × 2.5 mL PAXgene RNA tube. Tubes were shipped overnight to the Advanced Research and Diagnostic Laboratory at the University of Minnesota where processing occurred (within 48 hours of field collection time). DNA methylation profiles were obtained using the Infinium Methylation EPIC BeadChip with samples randomized across plates by key demographic variables, such as age, cohort, sex, education, and race^16^.

DNA methylation data preprocessing and quality control were performed in the R programming language with the R package *minfi* by the Health and Retirement Study^17,18^. Beta methylation values were calculated from the *minfi* RGChannelSet object. Methylation probes (3.4%, n=29,431 out of 866,091) were cut for failing detection P check (>0.01 detection P); detection P was then recalculated for samples and n=58 were removed (>0.05 detection P). Sex mismatched samples were also removed. This left 4,018 high-quality samples for analysis. Finally, missing beta methylation values were imputed with the mean beta methylation values of the given probe across all samples with present values^18^.

### DNA methylation clocks and age acceleration

The Health and Retirement Study released epigenetic clocks calculated in R with coefficients posted alongside the accompanying manuscript of each epigenetic clock^18^. The clocks can be categorized as phenotypic (capturing aging related outcomes) or chronologic (predicting traditional age). As our primary predictor, we prioritized the following phenotypic clocks: GrimAge, methylation pace of aging (DunedinPoAM38), and the Levine clock. The GrimAge clock incorporates biomarkers of physiological stress and DNA methylation based estimation of smoking pack-years to predict outcomes such as time to death, heart disease, and other age-related outcomes^19^. DunedinPoAM38 was developed using 18 biomarkers of rate of aging in various bodily systems (e.g., cardiovascular, renal, pulmonary systems)^20^. The Levine clock, based on whole blood using 513 CpGs, predicts several clinical outcomes such as mortality, cancer, and physical function^21^. Sensitivity analyses include three other chronological epigenetic clocks, described in **Supplemental Materials**.

For each clock, epigenetic age acceleration (i.e., age accelerated residuals) was calculated using linear models and by regressing each DNA methylation clock (Y) on participant’s chronological age (X) at wave 2016. Residual values greater than zero reflected epigenetic age acceleration and residual values less than or equal to zero reflected epigenetic age at or less than chronological age. Age acceleration residual measures were dichotomized at the zero (the mean) for interpretability purposes. We included continuous measure of age acceleration as sensitivity analyses of the joint associations of age acceleration and education on dementia and cognitive impairment, non-dementia.

### Cognitive status measures

We used a multidimensional measure of cognitive functioning based on a telephone-screening instrument: Telephone Interview for Cognitive Status^10^. Domains assessed using this measure include memory, mental status, abstract reasoning, fluid reasoning, vocabulary, dementia, and numeracy. In 2009, Langa, Kabeto, and Weir developed an approach for defining dementia and cognitively impaired non-dementia^26^ A team of dementia experts clinically validated this method using equipercentile equating against the Aging, Demographics, and Memory Study (ADAMS). The ADAMS study is a sub-sample of the Health and Retirement Study who received a more extensive neurological battery^27^. For self-respondents, the score consists of overall cognitive test performance. No proxy-rated respondents are included in these analyses. The cut points for this method reflect the prevalence of dementia or cognitive impairment to the expected population prevalence from the ADAMS study. All cognitive measures for these analyses were assessed in 2016 and taken from the imputed cognition researcher contribution data set^28^. A score from 0 to 6 is categorized as dementia, 7 to 11 is categorized as cognitive impaired non-dementia, and 12 to 27 is categorized as normal cognition^26^. We explored sensitivity models using the Power’s dementia classification – an expert-defined algorithmic dementia categorization designed to reduce bias misclassification of dementia for racial disparities research^29^.

### Covariate assessments

Self-reported educational attainment (years of school; categorized as “low education” for those with ≤12 years of education, and “high education” for those with >12 years of education), race/ethnicity (non-Hispanic White, non-Hispanic Black, Hispanic), and sex (0=female, 1=male) were measured at a participant’s initial exam. Chronological age (years) was assessed at the 2016 exam. Cell type proportions were estimated from a complete blood count. We included percent granulocytes and percent monocytes as precision variables in our models. Percent lymphocytes was not included, as the sum of percent granulocytes, monocytes, and lymphocytes was 100%.

### Four-level exposure variable

We created a four-level categorical variable consisting of all possible combinations between two dichotomized variables: education (≤ 12 years of education vs > 12 years of education) and epigenetic age acceleration (no age acceleration represented by an age acceleration residual value ≤ 0 vs accelerated aging with an age acceleration residual >0). Our four-level primary predictor represents the joint association of epigenetic clock and education. We organized the categories in the following way: a reference category representing the absence of both exposures (high education and age acceleration residuals ≤ 0); a second category denoting accelerated epigenetic aging (age acceleration residuals > 0 and high education); a third category denoting low education (≤ 12 years of education and age acceleration residuals ≤ 0); and a fourth category representing the presence of both exposures (≤ 12 years of education and age acceleration residuals > 0).

### Survey weights

We used survey weights for the Health and Retirement Study genetic sample from 2016 to generalize results to the Health and Retirement Study. We used these sample weights for descriptive statistics and all subsequent models.

### Statistical analysis

To properly account for the complex sampling design of the survey methodology and estimate weighted percentages, data were analyzed using the statistical software Stata 14^30^. Participants were included with complete data on DNA methylation clocks, covariates, cognitive status, and sample weight variables. We described covariate distributions using mean and standard deviation for continuous covariates and number and frequency for categorical covariates. We compared the distributions of covariates among included and excluded participants using t-tests for continuous covariates and chi-square tests for categorical covariates.

We calculated bivariate descriptive statistics on the combined sample and by cognitive status weighted by the 2016 genetic sample survey weights. We also tested the multivariable associations of each DNA methylation age acceleration method and education, adjusting for selected covariates, on either dementia or cognitive impairment non-dementia versus a reference category of normal cognition. These logistic models we weighted by the 2016 genetic sample survey weights.

We provide a conceptual model of our statistical analysis in **Figure 1**. We assessed additive interaction for all epigenetic clocks using impaired cognition as the outcome (i.e., dementia, cognitive impairment non-dementia, or Power’s dementia) relative to normal cognition, and we employed multivariable logistic regression to control for our main covariates of interest. Our base models included chronological age, self-reported race/ethnicity, sex, proportion granulocytes, proportion monocytes, and education as predictors. Subsequent models added dichotomized age acceleration residuals and an interaction between education and age acceleration. All models account for survey weights using Stata statistical software version 14^11^. We report odds ratios and 95% confidence intervals for interpreting effect estimates.

**Figure 1.**
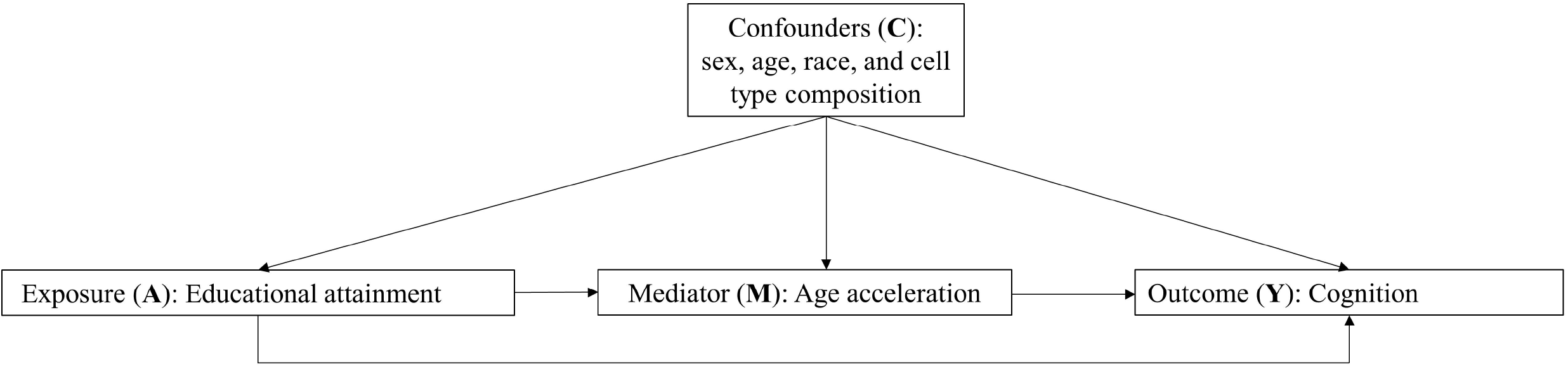
A heuristic model of our conceptual framework with exposure A (educational attainment), mediator M (age acceleration), outcome Y (cognition), and confounders C (sex, age, race, and cell type composition).

### Measures of interaction in the additive scale

We determined suitability to test for an additive interaction model of the joint association between an age acceleration measure (e.g. GrimAge acceleration, Levine age acceleration, etc.) and education if the sum of the odds ratio for the single exposed groups (OR_01_ + OR_10_, where OR_01_ is the association of DNA methylation age acceleration in those with educational attainment >12 years and OR_10_ is the association of educational attainment ≤12 in those with no DNA methylation age acceleration) was less than the odds ratio for the double exposed group (OR_11_, those with DNA methylation age acceleration and education ≤12 years). If these terms were satisfied, we would then calculate three measures of additive interaction for each age acceleration by education interaction: 1) the excess risk due to interaction, 2) the attributable proportion due to interaction, and 3) the synergy index. We used the delta method to calculate the corresponding 95% confidence intervals for these estimations^31^. In the context of our research, the excess risk due to interaction can be understood as the excess risk by which the interaction between age acceleration and ≤12 years of education exceed the addition of the isolated effects of both categories, that is, over and above their individual associations. The null value for the excess risk due to interaction is zero, a positive value is suggestive of synergism or interaction between the two variables and a negative value denotes antagonism. The attributable proportion due to interaction can be interpreted as the proportion of the excess risk in relation to cognitive impairment that is attributable to the interaction effect of epigenetic age acceleration and education ≤12 years; the null value for this measure is also zero^31^. Finally, the synergy index represents the ratio between the joint association of age acceleration and ≤12 years of education, divided by the isolated association of each of these two variables; a ratio of 1 represents the null value, and a ratio >1 is suggestive of superadditivity or synergism.

### Four-way mediation interaction decomposition analysis

After exploring the interaction effect between age acceleration and ≤12 years of education, we sought to understand whether epigenetic age acceleration mediates the association of education on cognitive impairment, while our mediation model accommodated for potential interaction effects between exposure and mediator. We decomposed this mediation-interaction analysis using the four-way decomposition method to estimate 1) the controlled direct effect of education, which indicates the association of education on cognitive impairment without age acceleration; 2) the interaction reference, which indicates the effect of the additive interaction that operates when age acceleration is present in those with > 12 years of education^32^. We also estimated 3) the interaction mediation, which represents the additive interaction that operates only if education has an effect on age acceleration at different levels of educational attainment; and 4) the pure indirect effect, or the effect of age acceleration on cognition in those with >12 years of education. The sum of these four components equate to the total effect of educational attainment on cognitive impairment. We used these four measures to calculate the percentage attributable to each effect, and employed the delta method to calculate associated confidence intervals for each estimation. Code to replicate these analyses is available at https://github.com/bakulskilab.

## RESULTS

### Study sample descriptive statistics

Of the 4,018 participants with DNA methylation measures, 3,724 had complete data for these analyses (**Figure 2**). Included and excluded participants were similar on most covariates, except self-reported race/ethnicity, sex, age, and cell type proportions (**Supplemental Table 1**). The analytic sample was predominantly non-Hispanic white (80.2%), female (53.9%), with an education >12 years (56.5%). The sample was 68.6 years on average. In bivariate descriptive statistics, the group with 12 or fewer years of education had higher weighted prevalence of dementia and cognitive impairment than the group with more than 12 years of education (**Table 1**).

**Figure 2.**
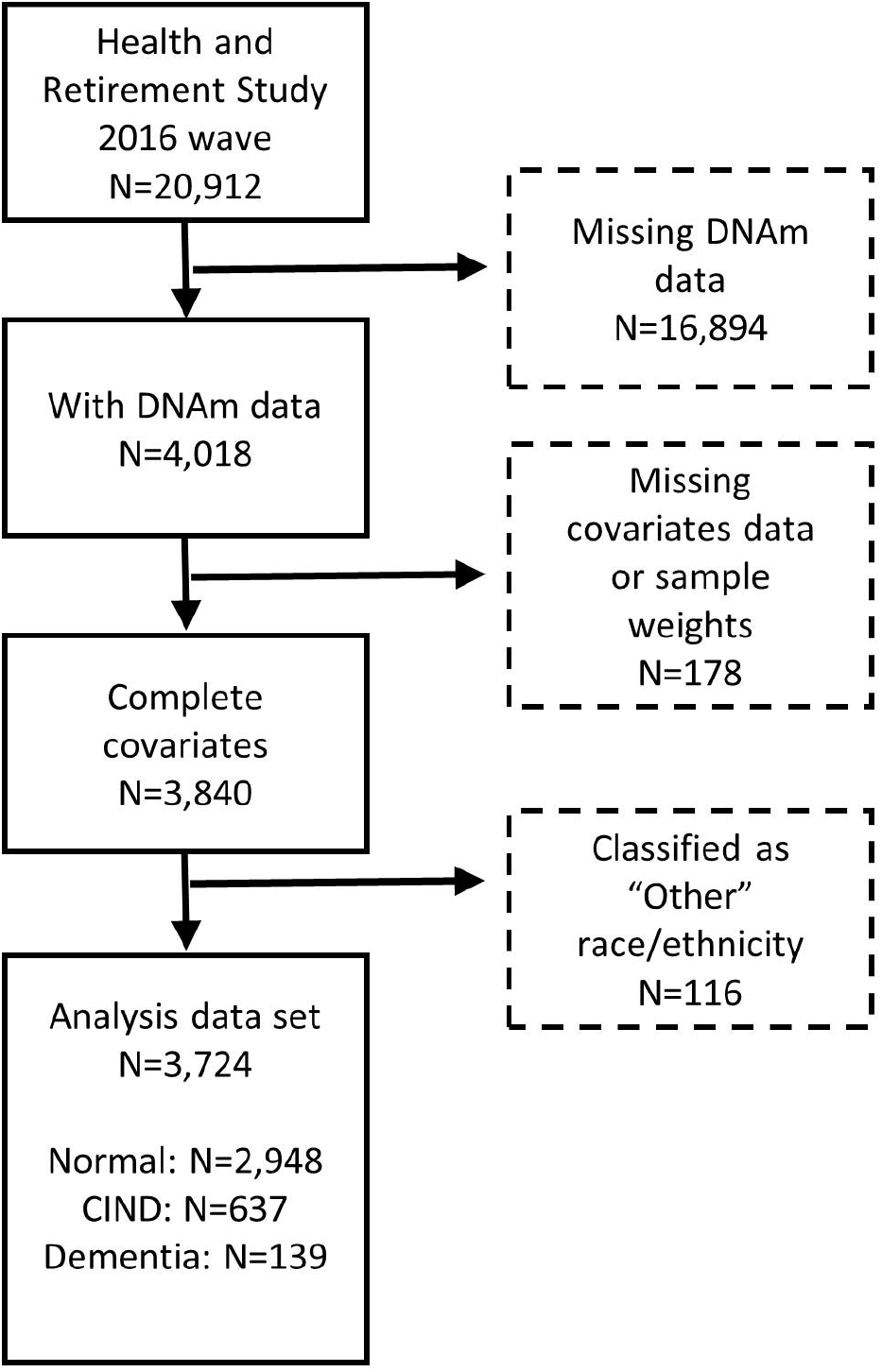
Analysis sample flow chart, Health and Retirement Study, 2016 wave. DNAm: DNA methylation; ClND: cognitively impaired, non-dementia. Analysis data set contains self-reported non-Hispanic white, non-Hispanic black, and Hispanic participants in the 2016 DNA methylation sample of the Health and Retirement Study.

**Table 1:** Weighted distribution of analytic sample covariates by cognitive status in the Health and Retirement Study, 2016 wave

|  | **Overall**<br>**N = 3,724** |  | **Normal cognition**<br>**N = 2,948** |  | **Cognitive Impairment, non-**<br>**dementia**<br>**N = 637** |  | **Dementia**<br>**N = 139** |  |
| --- | --- | --- | --- | --- | --- | --- | --- | --- |
|  | **Col%** | **95%CI** | **Col%** | **95%CI** | **Col%** | **95%CI** | **Col %** | **95%CI** |
| **Race** |  |  |  |  |  |  |  |  |
| Non-Hispanic White (n=2,724) | 80.2 | [78.8,81.5] | 83.7 | [82.3,85.1] | 65 | [60.4,69.3] | 49.7 | [39.9,59.6] |
| Non-Hispanic Black (n=628) | 10.5 | [9.5,11.6] | 8.4 | [7.4,9.5] | 20.3 | [16.7,24.5] | 24 | [17.0,32.7] |
| Hispanic (n=526) | 9.3 | [8.4,10.4] | 7.8 | [6.8,9.0] | 14.7 | [11.9,18.0] | 26.3 | [18.4,36.1] |
| **Sex** |  |  |  |  |  |  |  |  |
| Female (n=2,149) | 53.9 | [51.9,55.9] | 54.2 | [52.0,56.5] | 51.2 | [46.4,56.0] | 56.5 | [46.5,66.0] |
| Male (n=1,575) | 46.1 | [44.1,48.1] | 45.8 | [43.5,48.0] | 48.8 | [44.0,53.6] | 43.5 | [34.0,53.5] |
| **Education** |  |  |  |  |  |  |  |  |
| > 12 years (n=1,853) | 56.5 | [54.5,58.4] | 61.7 | [59.6,63.8] | 33.1 | [28.7,37.9] | 16.8 | [10.2,26.5] |
| ≤ 12 years (n=1,825) | 43.5 | [41.6,45.5] | 38.3 | [36.2,40.4] | 66.9 | [62.1,71.3] | 83.2 | [73.5,89.8] |
| **Chronological age** |  |  |  |  |  |  |  |  |
| Age in years | 68.6 | [68.3,69.0] | 67.5 | [67.2,67.9] | 73.4 | [72.4,74.4] | 77.6 | [75.3,79.9] |
| **Age accelerated residuals** | **Mean** | **95%CI** | **Mean** | **95%CI** | **Mean** | **95%CI** | **Mean** | **95%CI** |
| GrimAge | -0.2 | [-0.4,-0.1] | -0.6 | [-0.8,-0.4] | 1.4 | [0.9,1.9] | 1 | [-0.1,2.0] |
| DunedinPoAM38 | -0.4 | [-0.6,-0.1] | -0.6 | [-0.9,-0.4] | 1 | [0.4,1.6] | 0.8 | [-0.6,2.2] |
| Levine | 0 | [-0.3,0.3] | -0.2 | [-0.5,0.1] | 1.1 | [0.4,1.8] | 0 | [-1.3,1.2] |
| Horvath | 0 | [-0.2,0.3] | 0.1 | [-0.2,0.4] | -0.2 | [-0.8,0.5] | -0.4 | [-1.8,0.9] |
| Hannum | 0.1 | [-0.1,0.3] | 0.1 | [-0.2,0.3] | 0.5 | [0.0,0.9] | -0.6 | [-1.7,0.4] |
| Horvath - Skin | 0 | [-0.2,0.2] | 0 | [-0.2,0.2] | 0.2 | [-0.2,0.6] | -0.7 | [-1.6,0.3] |
| **Cell Type Proportions** | **Percent** | **95%CI** | **Percent** | **95%CI** | **Percent** | **95%CI** | **Percent** | **95%CI** |
| Granulocytes | 62 | [61.6,62.3] | 61.8 | [61.4,62.2] | 62.8 | [61.8,63.8] | 63.1 | [61.1,65.1] |
| Monocytes | 8.6 | [8.5,8.7] | 8.6 | [8.5,8.7] | 8.7 | [8.4,8.9] | 8.4 | [7.8,8.9] |
| Lymphocytes | 29.4 | [29.1,29.8] | 29.6 | [29.2,30.0] | 28.5 | [27.6,29.4] | 28.6 | [26.7,30.4] |

Note: Col%, Mean estimates and 95%CI represent statistics weighted using the 2016 genetic sample survey weights.

### Main associations of low education and high epigenetic age acceleration and cognitive status

We show results for effect estimates of dichotomous education and age acceleration in mutually adjusted, weighted logistic models in **Table 2**. All models were adjusted for chronological age, sex, race/ethnicity, and cell type proportions and weighted by the 2016 genetic sample weights. Low education (≤12 years of education) was significantly associated with dementia status (OR range 4.3-4.5, all P < 0.001) and also with cognitive impairment, non-dementia (OR range 2.4-2.6, all P < 0.001) (**Table 2**). High GrimAge acceleration was significantly associated with 1.63 times higher odds of cognitive impairment, non-dementia, relative to no age acceleration (95% CI, 1.3-2.1). No other dichotomous age acceleration variables were associated with dementia or cognitive impairment, non-dementia. We saw consistency in sensitivity models of continuous DNA methylation age acceleration (described in detail in **Supplemental Materials** and **Supplemental Table 2**).

**Table 2.** Weighted multivariable logistic regression results for mutually adjusted, education and dichotomized DNA methylation age acceleration residuals on dementia and cognitive impairment, non-dementia in the Health and Retirement Study, 2016 wave

|  | **Dementia vs normal cognition<sup>1</sup>** |  | **Cognitive impairment**<br>**non-dementia vs normal cognition<sup>2</sup>** |  |
| --- | --- | --- | --- | --- |
| **Age acceleration**<br>**calculation method<sup>+</sup>** | **OR** | **95%CI** | **OR** | **95%CI** |
| **GrimAge** |  |  |  |  |
| No age acceleration | 1 | - | 1 | - |
| Age acceleration | 1.49 | [0.90,2.49] | 1.63*** | [1.28,2.08] |
| High education | 1 | - | 1 | - |
| Low education | 4.32*** | [2.40,7.77] | 2.41*** | [1.91,3.04] |
| **DunedinPoAM38** |  |  |  |  |
| No age acceleration | 1 | - | 1 | - |
| Age acceleration | 1.12 | [0.67,1.88] | 1.19 | [0.93,1.52] |
| High education | 1 | - | 1 | - |
| Low education | 4.45*** | [2.49,7.95] | 2.50*** | [1.98,3.17] |
| **Levine** |  |  |  |  |
| No age acceleration | 1 | - | 1 | - |
| Age acceleration | 1.05 | [0.68,1.64] | 1.24 | [0.98,1.56] |
| High education | 1 | - | 1 | - |
| Low education | 4.49*** | [2.51,8.05] | 2.52*** | [1.99,3.18] |
| **Horvath** |  |  |  |  |
| No age acceleration | 1 | - | 1 | - |
| Age acceleration | 1.04 | [0.67,1.59] | 0.92 | [0.74,1.15] |
| High education | 1 | - | 1 | - |
| Low education | 4.50*** | [2.51,8.08] | 2.55*** | [2.01,3.22] |
| **Hannum** |  |  |  |  |
| No age acceleration | 1 | - | 1 | - |
| Age acceleration | 0.81 | [0.51,1.29] | 1.23 | [0.98,1.55] |
| High education | 1 | - | 1 | - |
| Low education | 4.51<sup>***</sup> | [2.51,8.10] | 2.53<sup>***</sup> | [2.00,3.20] |
| **Horvath-Skin** |  |  |  |  |
| No age acceleration | 1 | - | 1 | - |
| Age acceleration | 0.98 | [0.94,1.02] | 1.09 | [0.87,1.36] |
| High education | 1 | - | 1 | - |
| Low education | 4.37<sup>***</sup> | [2.41,7.94] | 2.54<sup>***</sup> | [2.01,3.21] |

Exponentiated Coefficients (Odds-Ratios); 95% confidence intervals in brackets
High education: > 12 years; low education ≤ 12 years
Age acceleration: >0 age acceleration residual; No age acceleration: ≤0 age acceleration residual
<sup>+</sup> Models adjusted for sex, chronological age, self-reported race/ethnicity (non-Hispanic White used as reference group), percent of granulocytes, and percent of monocytes
<sup>1</sup> Dementia vs normal cognition (Langa-Weir classification), n=3,087
<sup>2</sup> Cognitive impairment, non-dementia vs normal cognition (Langa-Weir classification), n=3,585
$p < 0.05$ , <sup>\*\*</sup> $p < 0.01$ , <sup>\*\*\*</sup> $p < 0.001$

### Interaction effects in the additive scale

We observed statistically significant interactions using all methods of epigenetic clock calculations between age deceleration and low education as well as age > and low education on both dementia and cognitive impairment, non-dementia compared to normal cognition, with no age acceleration and high education as the reference group (all *P*<0.05, **Table 3**). We additionally observed significant interactions between age acceleration and high education on cognitive impairment, non-dementia for both the GrimAge and DunedinPoAM38 epigenetic clock methods. For example, relative to the unexposed (those with no age acceleration and high education), participants with high GrimAge acceleration (and high education) had 1.5 times higher odds of dementia (95% CI, 0.4, 4.8), participants with low education (and no GrimAge acceleration) had 4.2 times higher odds of dementia (95% CI, 2.0-9.1), while participants with both low education and high GrimAge acceleration had 6.4 times higher odds of dementia (95% CI, 2.8-14.8).

**Table 3.** Weighted multivariable logistic regression additive interaction results for education and dichotomized DNA methylation age acceleration on dementia and impaired cognition, non-dementia in the Health and Retirement Study, 2016 wave

|  | Dementia vs normal cognition<sup>1</sup> |  |  |  |  |  | Cognitive impairment, non-dementia vs normal cognition<sup>2</sup> |  |  |  |  |
| --- | --- | --- | --- | --- | --- | --- | --- | --- | --- | --- | --- |
|  | No age acceleration + High education (reference) | Age acceleration + High education | No age acceleration + Low Education | Age acceleration + Low Education | Additivity assumption satisfied? |  | No age acceleration + High education (reference) | Age acceleration + High education | No age acceleration + Low Education | Age acceleration + Low education | Additivity assumption satisfied? |
| Age acceleration calculation method+ | OR<sub>00</sub> | OR<sub>01</sub> | OR<sub>10</sub> | OR<sub>11</sub> | OR<sub>11</sub> > OR<sub>01</sub> + OR<sub>10</sub> |  | OR<sub>00</sub> | OR<sub>01</sub> | OR<sub>10</sub> | OR<sub>11</sub> | OR<sub>11</sub> > OR<sub>01</sub> + OR<sub>10</sub> |
| **GrimAge** |  |  |  |  |  |  |  |  |  |  |  |
|  | 1 | 1.5 | 4.2*** | 6.4*** | Y |  | 1 | 2.0*** | 2.8*** | 4.1*** | N |
|  | - | [0.4,4.8] | [2.0,9.1] | [2.8,14.8] |  |  | - | [1.3,2.9] | [2.0,3.9] | [2.9,5.7] |  |
| **DunedinPoAM3 8** |  |  |  |  |  |  |  |  |  |  |  |
|  | 1 | 0.5 | 2.9** | 4.0*** | Y |  | 1 | 1.5* | 3.0*** | 3.1*** | N |
|  | - | [0.1,2.0] | [1.5,5.9] | [1.9,8.5] |  |  | - | [1.0,2.1] | [2.1,4.1] | [2.2,4.4] |  |
| **Levine** |  |  |  |  |  |  |  |  |  |  |  |
|  | 1 | 0.7 | 3.6** | 4.2*** | N |  | 1 | 1.3 | 2.6*** | 3.1*** | N |
|  | - | [0.2,2.2] | [1.6,7.9] | [1.9,9.4] |  |  | - | [0.9,1.8] | [1.9,3.6] | [2.3,4.4] |  |
| **Horvath** |  |  |  |  |  |  |  |  |  |  |  |
|  | 1 | 1.7 | 6.6*** | 6.0*** | N |  | 1 | 0.9 | 2.5*** | 2.3*** | N |
|  | - | [0.5,5.6] | [2.5,17.0] | [2.3,15.8] |  |  | - | [0.6,1.3] | [1.8,3.4] | [1.7,3.2] |  |
| **Hannum** |  |  |  |  |  |  |  |  |  |  |  |
|  | 1 | 1.2 | 5.5*** | 4.1** | N |  | 1 | 1.3 | 2.6*** | 3.1*** | N |
|  | - | [0.4,3.6] | [2.2,13.9] | [1.6,10.6] |  |  | - | [0.9,1.8] | [1.9,3.6] | [2.3,4.4] |  |
| **Horvath - Skin** |  |  |  |  |  |  |  |  |  |  |  |
|  | 1 | 0.5 | 3.2** | 3.4** | N |  | 1 | 1.2 | 2.7*** | 2.8*** | N |
|  | - | [0.2,1.4] | [1.4,7.0] | [1.5,7.5] |  |  | - | [0.8,1.7] | [2.0,3.8] | [2.0,3.9] |  |

Exponentiated Coefficients (Odds-Ratios); 95% confidence intervals in brackets
High education: > 12 years; low education ≤ 12 years
<sup>+</sup> Models adjusted for sex, chronological age, self-reported race/ethnicity (non-Hispanic White used as reference group), percent of granulocytes, and percent of monocytes
<sup>1</sup> Dementia vs normal cognition (Langa-Weir classification), n=3,087
<sup>2</sup> Cognitive impairment, non-dementia vs normal cognition (Langa-Weir classification), n=3,585
OR<sub>01</sub> is the effect of high DNA methylation age acceleration in those with educational attainment >12 years and OR<sub>10</sub> is the effect of educational attainment ≤12 in those with no DNA methylation age acceleration
\* $p < 0.05$ , \*\* $p < 0.01$ , \*\*\* $p < 0.001$

We determined if these associations were greater than additive by comparing the odds ratio in the double exposed group to the sum of the odds ratios in the single exposed groups. We observed greater than additive associations (OR_11_ > OR_01_ + OR_11_) in the models for GrimAge acceleration and the DunedinPoAM38 on dementia (**Table 3**). That is, for the GrimAge acceleration associations 6.4 > 1.5 + 4.2 and for the DunedinPoAM38 age acceleration residual associations 4.0 > 0.5 + 2.9 (**Table 3**). We performed four-way mediation-interaction effect decomposition on both these models.

The excess risk due to interaction between education and GrimAge acceleration was estimated to be 1.7 (95%CI, -1.2, 4.6), meaning the odds of dementia among participants with both exposures exceeds what one would expect if the association of age acceleration and low education were additive. Similarly, the attributable proportion was 0.3 (95%CI, -0.1, 0.6), suggesting that nearly 30% of the excess risk in the double exposed group can be explained by the interaction between the two exposures (low education and age acceleration). Finally, a synergy index of 1.5 (95%CI, 0.7, 2.7) indicates the presence of interaction between the two factors.

For DunedinPoAM38 acceleration, the excess risk due to interaction with education was estimated to be 1.5 (95%CI, -0.4, 3.5), meaning the odds of dementia among participants with both exposures exceeds what one would expect if the association of age acceleration and low education were additive. Similarly, the attributable proportion was 0.4 (95%CI, -0.0, 0.9), suggesting that nearly 40% of the excess risk in the double exposed group can be explained by the interaction between the two exposures (low education and age acceleration). Finally, a synergy index of two (95%CI, 0.7, 6.1) indicates the presence of interaction between the two factors. Of note is that for those with age acceleration and high education, the odds of dementia was less than one, though not significantly different from one. This may introduce a potential violation of the additivity assumptions.

### Four-way mediation-interaction effect decomposition

We performed a four-way effect decomposition analysis for education and age acceleration for the GrimAge and DunedinPoAM38 methods on dementia. We set continuous covariates at the mean (age, percent granulocytes, and percent monocytes) and sex as male. We separately estimated four-way mediation-interaction effects for each of the three self-reported race/ethnicities (non-Hispanic white, non-Hispanic Black, and Hispanic). For GrimAge acceleration, this mediation-interaction decomposition analysis suggests that most of the association of education on cognition is due to the controlled direct effect. The percent attributable to the controlled direct effect of education on dementia status was 74.6% (95%CI, 40.6-108.7) for the non-Hispanic White reference group, 69.5% (95%CI, 29.9-109.1) for the non-Hispanic Black group, and 74.4% (95%CI, 40.1-108.7) for the Hispanic group (**Table 4)**. Additionally, we found that the percent of the association of education on cognition that is mediated through GrimAge acceleration is around 8.4% (95%CI, -1.1-17.9) for the non-Hispanic White group; 6.0% (95%CI, -0.2-12.2) for the non-Hispanic Black group; and 8.4% (95%CI, -1.0-17.7) for the Hispanic group. We calculated these estimates by adding the interaction mediation and the pure indirect effects. We found similar results for DunedinPoAM38 age acceleration in the percent attributable to the controlled direct effect of education on dementia status. The percent of the association of education on cognition that is mediated through DunedinPoAM38 acceleration is 5.0% (95%CI,-3.5-13.5) for the non-Hispanic White group, 3.6% (95%CI, -2.1-9.3) for the non-Hispanic Black group, and 4.8% (95%CI, -3.1-12.7) for the Hispanic group. Sensitivity models using the Power’s dementia classification can be found in **Supplemental Materials** and **Supplemental Tables 2, 3**, and **4**.

**Table 4:** Four-way effect decomposition results for high education and no DNA methylation age acceleration based on the GrimAge and DunedinPoAM38 method on dementia in the Health and Retirement Study 2016 wave

|  | GrimAge |  |  |  | DunedinPoAM38 |  |  |  |
| --- | --- | --- | --- | --- | --- | --- | --- | --- |
|  | non-Hispanic White |  |  |  | non-Hispanic White |  |  |  |
|  | Excess effect<br>on OR scale | 95%CI | % Attributable | 95%CI | Excess effect<br>on OR scale | 95%CI | % Attributable | 95%CI |
| **Controlled Direct Effect** | 3.2 | [-0.0,6.5] | 74.6% | [40.6,108.7] | 1.9 | [-0.1,4.0] | 73.4% | [38.6,108.2] |
| **Interaction Reference** | 0.7 | [-0.5,2.0] | 17.0% | [-8.9,42.9] | 0.6 | [-0.2,1.3] | 21.7% | [-1.8,16.4] |
| **Interaction Mediation** | 0.3 | [-0.2,0.8] | 6.6% | [-3.5,16.7] | 0.2 | [-0.1,0.4] | 7.3% | [-1.8,16.4] |
| **Pure Indirect Effect** | 0.1 | [-0.2,0.4] | 1.8% | [-4.3,7.9] | -0.1 | [-0.1,0.0] | -2.3% | [-6.8,2.2] |
| **Total** | 4.4 | [0.4,8.3] |  |  | 2.6 | [0.4,4.9] |  |  |
| **Portion Mediated** |  |  | 8.4% | [-1.1,17.9] |  |  | 5.0% | [-3.5,13.5] |
| **Portion due to Interaction** |  |  | 23.6% | [-12.4,59.6] |  |  | 28.9% | [-7.3,65.2] |
|  | non-Hispanic Black |  |  |  | non-Hispanic Black |  |  |  |
|  | Excess effect<br>on OR scale | 95%CI | % Attributable | 95%CI | Excess effect<br>on OR scale | 95%CI | % Attributable | 95%CI |
| **Controlled Direct Effect** | 3.2 | [-0.0,6.5] | 69.5% | [29.9, 109.1] | 1.9 | [-0.1,4.0] | 63.6% | [23,104.2] |
| **Interaction Reference** | 1.1 | [-0.8,3.1] | 24.5% | [-10.1,59.1] | 1.0 | [-0.3,2.3] | 32.8% | [-2.9,68.4] |
| **Interaction Mediation** | 0.2 | [-0.2,0.6] | 4.7% | [-1.9,11.3] | 0.2 | [-0.0,0.4] | 5.3% | [-0.5,11.1] |
| **Pure Indirect Effect** | 0.1 | [-0.2,0.3] | 1.3% | [-3.1,5.7] | -0.1 | [-0.1,0.0] | -1.7% | [-4.9,1.5] |
| **Total** | 4.7 | [0.5,8.8] |  |  | 3.0 | [0.6,5.5] |  |  |
| **Portion Mediated** |  |  | 6.0% | [-0.2,12.2] |  |  | 3.6% | [-2.1,9.3] |
| **Portion due to Interaction** |  |  | 29.2% | [-12.0,70.4] |  |  | 38.1% | [-3.3,79.5] |
|  | Hispanic |  |  |  | Hispanic |  |  |  |
|  | Excess effect<br>on OR scale | 95%CI | % Attributable | 95%CI | Excess effect<br>on OR scale | 95%CI | % Attributable | 95%CI |
| **Controlled Direct Effect** | 3.2 | [-0.0,6.5] | 74.4% | [40.1,108.7] | 1.9 | [-0.1,4.0] | 69.9% | [32.6,107.2] |
| **Interaction Reference** | 0.8 | [-0.6,2.1] | 17.2% | [-9.0,43.5] | 0.7 | [-0.2,1.6] | 25.3% | [-4.9,55.6] |
| **Interaction Mediation** | 0.3 | [-0.2,0.8] | 6.6% | [-3.4,16.6] | 0.2 | [-0.1,0.4] | 7% | [-1.4,15.3] |
| **Pure Indirect Effect** | 0.1 | [-0.2,0.4] | 1.8% | [-4.3,7.8] | -0.1 | [-0.1,0.0] | -2.2% | [-6.5,2.1] |
| **Total** | 4.4 | [0.5,8.3] |  |  | 2.8 | [0.5,5.1] |  |  |
| **Portion Mediated** |  |  | 8.4% | [-10,17.7] |  |  | 4.8% | [-3.1,12.7] |
| Portion due to Interaction |  |  | 23.8% | [-12.4,60.1] |  |  | 32.3% | [-6.3,70.9] |

Models are adjusted (self-reported race/ethnicity as noted, male, of average age, average percent of granulocytes and monocytes) and weighted to be representative of the Health and Retirement Study.

## DISCUSSION

Inequities in early education can affect biological aging and perpetuate disparities in cognition later in life. In a large, nationally representative sample of older adults in the United States, we characterized the relationships between epigenetic age acceleration measured in blood and education with dementia and cognitive impairment, non-dementia. We observed a 1.6 times increase in the odds of cognitive impairment, non-dementia in those with high DNA methylation age acceleration characterized using the GrimAge method relative to those with no DNA methylation age acceleration, after adjustment. Further, we saw a more than four times increase in the odds of dementia and a roughly 2.5 times increase in the odds of cognitive impairment, non-dementia in those with low education (≤12 years) compared to those with high education, after accounting for age acceleration. We observed interaction effects in the additive scale between low education and age acceleration using the GrimAge and DunedinPoAM38 methods on the odds of dementia. This includes a 6.4 times (95%CI, 2.8-14.8) higher odds of dementia in those with low education and high GrimAge acceleration and a 4 times (95%CI, 1.9-8.5) higher odds of dementia in those with low education and high DunedinPoAM38 age acceleration relative to the unexposed. We also observed that 6-8% of the proportion of the association of education on dementia is mediated through GrimAge acceleration while 3.6-5% of the association of education on dementia may be mediated through DunedinPoAM38 age acceleration. In sensitivity analyses using the Power’s dementia algorithm to categorize dementia with greater attention to defining the outcome to elucidate racial disparities, we observed the association of education on cognition to be mediated at a much higher percent: 11-16.3%. Though there is need for additional research, these findings suggest the association of DNA methylation age acceleration using methods sensitive to phenotypes (i.e. GrimAge, Levine, and DunedinPoAM38) is small, but present with prevalent dementia. Our study further suggests DNA methylation age acceleration may interact with education in an additive manner in its association on dementia and impaired cognition.

To fully understand the efficacy of epigenetic age acceleration as a biomarker of dementia, it is critical to understand how socioeconomic factors including education modify observed associations between epigenetic aging and dementia across different race/ethnic populations. Epigenetic studies on minoritized social groups have been underpowered, and unfortunately, work on these same processes in Hispanic populations has been almost nonexistent—especially in nationally representative samples^48-50^. Our study has taken steps to characterize associations between education, DNA methylation age acceleration, and cognition within groups racialized as non-Hispanic Black, non-Hispanic White, and Hispanic as well as examining a dementia outcome variable sensitive to race/ethnic disparities.

Our findings extend prior research linking biological aging and memory decline in later life from smaller, White European and New Zealand samples^20,21,33^. One recent study, using the same DNA methylation sub-sample of the Health and Retirement Study found that participants with lower SES – defined as a combination of education and wealth variables - had lower memory performance, faster decline and exhibited accelerated biological aging (SES effect size associations (β) ranged from 0.08 to 0.41)^34^. This group further found that biological aging using DNA methylation accounted for 4-27% of the SES-memory gradient in White respondents, but there was minimal evidence of mediation in the Black or Latinx participants. This study examined three of the same epigenetic age acceleration variables as in our study: GrimAge, DunedinPoAM38, and the Levine methods. Consistent with their findings that accelerated biological aging was associated with lower memory performance, we did find an association between high GrimAge acceleration and cognitive impairment, non-dementia and between continuous DNA methylation age acceleration residuals and dementia, but we did not see associations between any of the other age acceleration methods and impaired cognition. Counter to their conclusion, though we did find some evidence for mediation, the percentage was small (6-8.4%) and the confidence intervals contained the null value of 0%. This value also did not appreciably change when using different racialized groups as reference categories. We did not investigate sex/gender differences within the context of race/ethnic groups due to sample size concerns.

One potential limitation of our study is that DNA methylation was measured in venous blood samples. While brain tissue may appear to be the neurodegenerative disease epigenetic “gold standard”, the postmortem acquisition timing, scarcity of confounder/exposure data, and limited sample sizes with lack of replication potential present many limitations^35^.Postmortem samples may reflect epigenetic consequence of disease rather than cause; there may be bias in biological signals due competing causes of death^36^, tissue pH ^37^, or pre-mortem agonal state^38^. Peripheral blood with minimally invasive collection is the most abundant sample type for epidemiologic and clinical research. DNA methylation measured in peripheral tissues are associated with mortality^39-41^, morbidity^14,25,39,41-43^, and healthy aging^21,23,44,45^. Importantly, peripheral DNA methylation is linked to impaired cognition^11-13^, though much of this work has been on global or candidate gene DNA methylation. Epigenetic clocks representing accelerated aging have shown promise as potential biomarkers of dementia. Importantly, these age acceleration methods can be applied to DNA methylation obtained from whole blood, a relatively convenient and minimally invasive source for biomarkers.

The effect sizes we observed for different methods of DNA methylation age acceleration varied by method and in significance. While these methods of characterizing age acceleration are potentially useful as biomarkers of later life cognition, DNA methylation is measured imprecisely and these methods have varied reliability^46^. While many new and improved methods to assess DNA methylation biomarkers are being developed, our results only provide a beginning to the investigation of the utility of DNA methylation age acceleration and its relationship to education and cognition and definitive conclusions based on these preliminary results should be avoided.

## Conclusion

Alzheimer’s disease and its related dementias remain prevalent health burdens that are unevenly distributed in the population across socioeconomic factors. It is imperative to identify conveniently obtainable biomarkers of dementia that can clarify the biologic underpinnings of dementia as well as modifiable environmental or socioeconomic factors that contribute to disparities in dementia risks. Our results demonstrate that age acceleration measured using DNA methylation clock methods differ in their associations with dementia and cognitive impairment status. Although additional research is needed, our results suggest that education does not substantively mediate the association between age acceleration and dementia or cognitive impairment. We do find a potential for additive interaction between education and DNA methylation age acceleration in our sample from the United States nationally representative Health and Retirement Study. Our findings highlight the need for more, large, multi-ethnic, population-based epigenetic studies of dementia that account for socioeconomic factors including education to better understand the interplay of social disadvantage and the biological aging process.

## Data Availability

All data used are publicly available through the Health and Retirement Study.

https://hrs.isr.umich.edu/

## Data availability statement

All data used are publicly available through the Health and Retirement Study (https://hrs.isr.umich.edu/).

We thank the participants and staff of the Health and Retirement Study. We thank Brittni Delmaine for her editorial services and the Dr. Jennifer Smith lab at the University of Michigan School of Public Health for valuable suggestions in this analysis.

