## Supplemental for "Additive interaction and mediation-interaction decomposition: DNA methylation age acceleration, education, and cognitive impairment in the Health and Retirement Study"

426 Thompson St

Ann Arbor, MI, 48104

USA

**Keywords:** DNA methylation, Methylation Clocks, Age acceleration, Dementia, Disparities

**Abstract**

**Importance:** Dementia represents a significant and increasing public health burden. DNA methylation age acceleration may be associated with dementia and dementia risk factors, such as education, but investigating their impact on dementia is necessary.

**Objective:** To evaluate the association of educational attainment on dementia and cognitive impairment through DNA methylation age acceleration, while accommodating exposure-mediator interaction effects.

**Design:** In the 2016 Health and Retirement Study wave, we evaluated six epigenetic clocks, including GrimAge, with Langa-Weir classified dementia, cognitive impairment non-dementia, and normal cognition. Age acceleration was the residual between chronological age of participants and DNA methylation clock, dichotomized at zero. To understand the joint association of low education (≤12 years) and DNA methylation age acceleration in relation to cognitive impairment, we used weighted logistic regression and calculated interaction on the additive scale adjusting for chronological age, sex, race/ethnicity, and cell type composition. We performed four-way mediation and interaction decomposition analysis to estimate the: 1) controlled direct effect of education on cognition, 2) interaction reference, 3) interaction mediation, and 4) the pure indirect effect of DNA methylation age acceleration on cognition.

**Setting:** Analysis was conducted on a subsample of Health and Retirement Study participants in the 2016 Venous Blood Study (N=3,724).

**Results**: Both GrimAge acceleration (OR=1.6 95%CI 1.3 – 2.1) and low educational attainment (OR= 2.4 95%CI 1.9 – 3.0) were associated with higher odds of cognitive impairment, non-dementia in a mutually adjusted logistic model. We found additive interaction associations between low education and GrimAge acceleration on dementia. We observed that 6-8% of the association of education on dementia was mediated through GrimAge acceleration. While mediation effects were small, the portion of the association of education due to additive interaction with GrimAge acceleration was between 23.6 and 29.2%.

**Conclusions and Relevance:** Accelerated DNA methylation age was associated with increased odds of cognitive impairment and we observed more than additive interaction effects between education and age acceleration on dementia. These results support the interplay of social disadvantage and biological aging processes on impaired cognition.

**SUPPLEMENTAL METHODS**

*DNA methylation clocks and age acceleration*

In sensitivity analyses we examined an additional three chronologic epigenetic clocks. The Horvath clock is a multi-tissue clock developed from 8,000 samples. This clock incorporates 51 tissues and cell types and estimates DNA methylation based on 353 CpG sites^1,2^. The Hannum clock is a blood-based clock that uses 71 CpG sites. This clock is based on whole blood taken from 656 human samples ranging in age from 19 to 101^3^. The Horvath (Skin) clock is based on 391 CpGs and was developed to better measure the age of skin cell types such as keratinocytes as well as buccal cells, endothelial cells, and other sample types such as blood and saliva^4^.

**SUPPLEMENTAL RESULTS**

*Main associations of low education and high epigenetic age acceleration and cognitive status*

A one-year increase in GrimAge acceleration was associated with 1.06 times higher odds of dementia (95% CI, 1.0-1.1), relative to normal cognition. Age acceleration from the DunedinPoAM38, Levine, Horvath, Hannum, and Horvath skin methods were not associated with dementia, after adjusting for covariates and education. A one-year increase in GrimAge acceleration was also associated with 1.08 times higher odds of cognitive impairment non-dementia (95% CI, 1.0-1.1), relative to normal cognition. Age acceleration from the DunedinPoAM38, Levine, Hannum, and Horvath skin methods were significantly associated with cognitive impairment, non-dementia, after adjusting for the aforementioned covariates and education (**Supplemental** **Table 2**).

*Sensitivity models using the Power’s dementia classification^5^*

In our analysis sample of 3,724 participants, 3,694 also had a dementia classification using the Power’s dementia algorithm ^6^. Almost 94% (n=3,464, 93.8%) of the sample with a measurement for Power’s dementia were classified as normal cognition. The Power’s normal cognition classification largely overlapped with the Langa-Weir normal cognition category. There were 2,986 participants (80.8%) with concordant classifications (i.e. dementia/dementia or normal/normal) between the Langa-Weir and Power’s dementia algorithms. The majority of respondents classified as having cognitive impairment, non-dementia (n=499, 79.3%) were reclassified as having normal cognition using the Power’s algorithm. A small proportion of participants moved from normal cognition to dementia (n=23, 0.6%) or from dementia to normal cognition (n=56, 1.5%) when moving from the Langa-Weir to Power’s algorithm to classify dementia.

Results for effect estimates of dichotomous education and age acceleration in mutually adjusted, weighted logistic models are shown in **Supplemental** **Table 2**. Education was significantly associated with Power’s dementia classification, adjusting for sex, chronological age, self-reported race/ethnicity, percent of granulocytes, percent of monocytes, and age acceleration (all methods, **Supplemental Table 3**). Age acceleration using the DunedinPoAM38 method was associated with 1.7 times higher odds of dementia (95%CI, 1.2-2.5). No other methods of age acceleration were associated with dementia when dichotomized. When modeled as continuous, age acceleration residuals using the GrimAge and Levine methods were significantly associated with Power’s dementia, again in weighted logistic models adjusting for sex, chronological age, self-reported race/ethnicity, percent of granulocytes, percent of monocytes, and education (**Supplemental Table 2**).

DunedinPoAM38 acceleration showed more than additive interaction effects. Relative to the unexposed (those with low age acceleration and high education), participants with high DunedinPoAM38 age acceleration (and high education) had 1.1 times higher odds of dementia (95% CI, 0.6-2.1), participants with low education (and low DunedinPoAM38 age acceleration) had 1.9 times higher odds of dementia (95% CI, 1.1-3.3), while participants with both low education and high DunedinPoAM38 age acceleration had 2.0 times higher odds of dementia (95% CI,1.2-3.6). These weighted models were adjusted for sex, chronological age, self-reported race/ethnicity, percent of granulocytes, and percent of monocytes (**Supplemental** **Table 4)**.

For DunedinPoAM38 acceleration, the excess risk due to interaction with education was estimated to be 1. (95%CI, 0.1, 2.3), meaning the odds of dementia among participants with both exposures exceeds what one would expect if the association of age acceleration and low education were additive. Similarly, the attributable proportion was 0.4 (95%CI, 0.0, 0.7), suggesting that nearly 40% of the excess risk in the double exposed group can be explained by the interaction between the two exposures (low education and age acceleration). Finally, a synergy index of 2.7 (95%CI, 0.5, 13.5) indicates the presence of interaction between the two factors.

For DunedinPoAM38 acceleration, mediation-interaction decomposition analysis suggests that most of the association of education on cognition is due to the controlled direct effect. The percent attributable to the controlled direct effect of education on dementia status was 40.4% (95%CI: -13.1-94.0) for the non-Hispanic White group; 31.9% (95%CI: -17.3-81.2) for the non-Hispanic Black group; and 37.8% (95%CI: -14.7-90.2) for the Hispanic group (**Supplemental Table 5)**. Additionally, we found that the percent of the association of education on cognition mediated through DunedinPoAM38 acceleration is 16.2% (95%CI, 4.6-27.8) for the non-Hispanic White group; 11.0% (95%CI, 4.8-17.1) for the non-Hispanic Black group; and 15.3% (95%CI, 5.2-25.4) for the Hispanic group.

**Supplemental Table 1:** Distribution of included versus excluded samples in the Health and Retirement Study, wave 2016 DNA methylation sample.

|  | **Total** | **Included** | **Excluded** | **p-value^*^** |
| --- | --- | --- | --- | --- |
|  | (n=4,018) | (n=3,724) | (n=294) |  |
|  | **Col%** | **Col%** | **Col %** |  |
| **Cognitive Status** |  |  |  | 0.703 |
| Dementia | 3.8 | 3.7 | 4.4 |  |
| Cognitive impairment non-dementia | 17 | 17.1 | 15.6 |  |
| Normal cognition | 79.2 | 79.2 | 79.9 |  |
| **Self-reported race /ethnicity** |  |  |  | 0.001 |
| Non-Hispanic Black | 16.9 | 16.9 | 17.2 |  |
| Hispanic | 14.6 | 14.1 | 24.3 |  |
| Non-Hispanic White | 68.6 | 69 | 58.6 |  |
| **Sex** |  |  |  | 0.001 |
| Female | 58.5 | 57.7 | 68 |  |
| Male | 41.5 | 42.3 | 32 |  |
| **Education** |  |  |  | 0.786 |
| > 12 years | 50.6 | 50.5 | 51.4 |  |
| ≤ 12 years | 49.4 | 49.5 | 48.6 |  |
| **Chronological age** | **Mean** | **Mean** | **Mean** |  |
| Age in years | 69.4 | 70.1 | 60.9 | <0.001 |
| **Age accelerated residuals** | **Mean** | **Mean** | **Mean** |  |
| GrimAge | 0 | 0.01 | -0.14 | 0.617 |
| DunedinPoAM38 | 0 | -0.04 | 0.55 | 0.085 |
| Levine | 0 | -0.05 | 0.64 | 0.127 |
| Horvath | 0 | -0.02 | 0.29 | 0.447 |
| Hannum | 0 | 0.01 | -0.09 | 0.771 |
| Horvath - Skin | 0 | 0.04 | -0.48 | 0.041 |
| **Cell Type Proportions** | **Percent** | **Percent** | **Percent** |  |
| Granulocytes | 61.6 | 61.7 | 59.3 | <0.001 |
| Monocytes | 8.5 | 8.6 | 7.7 | <0.001 |
| Lymphocytes | 29.9 | 29.7 | 32.9 | <0.001 |

* P-values obtained from chi^2^ test for categorical variables, and T-test of independence with unequal variance for continuous variables

Note: Column percentages of included versus excluded samples and total Venous Blood Sample

of epigenetic accelerated aging in wave 2016 of the Health and Retirement Study

**Supplemental Table 2.** Weighted multivariable logistic regression results for mutually adjusted, education and continuous DNA methylation age acceleration residuals on dementia and cognitive impairment, non-dementia in the Health and Retirement Study, 2016 wave

|  | **Dementia vs normal cognition^1^** | | **Cognitive impairment**  **non-dementia vs normal cognition^2^** | | **Dementia vs normal cognition^3^** | |
| --- | --- | --- | --- | --- | --- | --- |
| **Age acceleration calculation method+** | **OR** | **95%CI** | **OR** | **95%CI** | **OR** | **95%CI** |
| **GrimAge** |  |  |  |  |  |  |
| Age acceleration | 1.06^*^ | [1.00,1.13] | 1.08^***^ | [1.04,1.11] | 1.06^*^ | [1.01,1.11] |
| High education | 1 | - | 1 | - | 1 | - |
| Low education | 4.08^***^ | [2.24,7.43] | 2.32^***^ | [1.83,2.94] | 1.81^**^ | [1.24,2.65] |
| **DunedinPoAM38** |  |  |  |  |  |  |
| Age acceleration | 1.02 | [0.98,1.06] | 1.02^*^ | [1.00,1.04] | 1.01 | [0.98,1.04] |
| High education | 1 | - | 1 | - | 1 | - |
| Low education | 4.30^***^ | [2.39,7.73] | 2.48^***^ | [1.96,3.14] | 1.87^**^ | [1.28,2.73] |
| **Levine** |  |  |  |  |  |  |
| Age acceleration | 1.00 | [0.98,1.03] | 1.02^**^ | [1.01,1.04] | 1.03^*^ | [1.01,1.05] |
| High education | 1 | - | 1 | - | 1 | - |
| Low education | 4.37^***^ | [2.41,7.95] | 2.53^***^ | [2.00,3.20] | 1.90^***^ | [1.30,2.78] |
| **Horvath** |  |  |  |  |  |  |
| Age acceleration | 1.01 | [0.98,1.04] | 1.00 | [0.98,1.01] | 1.00 | [0.98,1.03] |
| High education | 1 | - | 1 | - | 1 | - |
| Low education | 4.38^***^ | [2.41,7.98] | 2.54^***^ | [2.01,3.21] | 1.88^**^ | [1.29,2.75] |
| **Hannum** |  |  |  |  |  |  |
| Age acceleration | 0.99 | [0.95,1.03] | 1.02^*^ | [1.00,1.04] | 1.02 | [0.99,1.05] |
| High education | 1 | - | 1 | - | 1 | - |
| Low education | 4.38^***^ | [2.41,7.96] | 2.53^***^ | [2.00,3.20] | 1.88^**^ | [1.28,2.74] |
| **Horvath-Skin** |  |  |  |  |  |  |
| Age acceleration | 0.98 | [0.94,1.02] | 1.01 | [0.99,1.04] | 1.00 | [0.97,1.04] |
| High education | 1 | - | 1 | - | 1 | - |
| Low education | 4.37^***^ | [2.41,7.94] | 2.54^***^ | [2.01,3.21] | 1.88^**^ | [1.28,2.74] |

Exponentiated Coefficients (Odds-Ratios); 95% confidence intervals in brackets

High education: > 12 years; low education ≤ 12 years

^+^ Models adjusted for sex, chronological age, self-reported race/ethnicity (non-Hispanic White used as reference group), percent of granulocytes, and percent of monocytes

^1^ Dementia vs normal cognition (Langa-Weir classification), n=3,087

^2^ Cognitive impairment, non-dementia vs normal cognition (Langa-Weir classification), n=3,585

^3^ Power’s dementia classification vs normal cognition (Langa-Weir classification), n=3,694

^*^ *p* < 0.05, ^**^ *p* < 0.01, ^***^ *p* < 0.001

**Supplemental Table 3.** Weighted multivariable logistic regression results for mutually adjusted, education and dichotomized DNA methylation age acceleration residuals on Power’s algorithm defined dementia in the Health and Retirement Study, 2016 wave

|  | **Dementia vs normal cognition^1^** | |
| --- | --- | --- |
| **Age acceleration calculation method^+^** | **Odds ratio** | **95%CI** |
| **GrimAge** |  |  |
| No age acceleration | 1 | - |
| Age acceleration | 1.08 | [0.72,1.63] |
| High education | 1 | - |
| Low education | 1.87^**^ | [1.28,2.73] |
| **DunedinPoAM38** |  |  |
| No age acceleration | 1 | - |
| Age acceleration | 1.71^**^ | [1.15,2.54] |
| High education | 1 | - |
| Low education | 1.85^**^ | [1.26,2.70] |
| **Levine** |  |  |
| No age acceleration | 1 | - |
| Age acceleration | 1.31 | [0.91,1.89] |
| High education | 1 | - |
| Low education | 1.89^**^ | [1.29,2.76] |
| **Horvath** |  |  |
| No age acceleration | 1 | - |
| Age acceleration | 1.15 | [0.80,1.66] |
| High education | 1 | - |
| Low education | 1.88^**^ | [1.29,2.75] |
| **Hannum** |  |  |
| No age acceleration | 1 | - |
| Age acceleration | 1.25 | [0.84,1.84] |
| High education | 1 | - |
| Low education | 1.88^**^ | [1.29,2.75] |
| **Horvath-Skin** |  |  |
| No age acceleration | 1 | - |
| Age acceleration | 1.13 | [0.79,1.63] |
| High education | 1 | - |
| Low education | 1.88^**^ | [1.28,2.74] |

Exponentiated Coefficients (Odds-Ratios); 95% confidence intervals in brackets

High education: > 12 years; low education ≤ 12 years

Age acceleration: ≥0 age acceleration residual; No age acceleration: <0 age acceleration residual

^+^ Models adjusted for sex, chronological age, self-reported race/ethnicity (non-Hispanic White used as reference group), percent of granulocytes, and percent of monocytes

^1^ Dementia vs normal cognition (Power’s classification), n=3,694

^*^ *p* < 0.05, ^**^ *p* < 0.01, ^***^ *p* < 0.001

**Supplemental Table 4.** Multivariable logistic regression additive interaction results for education and DNA methylation age acceleration on Power’s algorithm dementia classification in the Health and Retirement Study, 2016 wave

|  | No age acceleration + High education | Age acceleration + High education | No age acceleration + Low Education | Age acceleration + Low education | Additivity assumption satisfied? |
| --- | --- | --- | --- | --- | --- |
| **Age acceleration calculation method^+^** | OR_00_ | OR_01_ | OR_10_ | OR_11_ | OR_11_ > OR_01_ + OR_10_ |
| **GrimAge** |  |  |  |  |  |
|  | 1 | 1.1 | 1.9^*^ | 2.0^*^ | N |
|  | - | [0.6,2.1] | [1.1,3.3] | [1.2,3.6] |  |
| **DunedinPoAM38** |  |  |  |  |  |
|  | 1 | 1.3 | 1.4 | 2.8^***^ | Y |
|  | - | [0.7,2.4] | [0.8,2.5] | [1.6,5.0] |  |
| **Levine** |  |  |  |  |  |
|  | 1 | 1.4 | 2.0^*^ | 2.5^**^ | N |
|  | - | [0.8,2.6] | [1.1,3.5] | [1.4,4.4] |  |
| **Horvath** |  |  |  |  |  |
|  | 1 | 1.1 | 1.8^*^ | 2.1^**^ | N |
|  | - | [0.6,2.0] | [1.0,3.0] | [1.2,3.6] |  |
| **Hannum** |  |  |  |  |  |
|  | 1 | 1.7 | 2.4^**^ | 2.5^**^ | N |
|  | - | [0.9,3.2] | [1.3,4.2] | [1.4,4.7] |  |
| **Horvath - Skin** |  |  |  |  |  |
|  | 1 | 1.1 | 1.8^*^ | 2.1^**^ | N |
|  | - | [0.6,2.0] | [1.1,3.1] | [1.2,3.6] |  |

Exponentiated Coefficients (Odds-Ratios); 95% confidence intervals in brackets

High education: > 12 years; low education ≤ 12 years

^+^ Models adjusted for sex, chronological age, self-reported race/ethnicity (non-Hispanic White used as reference group), percent of granulocytes, and percent of monocytes

^1^ Dementia vs normal cognition (Power’s classification), n=3,694

OR_01_ is the effect of high DNA methylation age acceleration in those with educational attainment >12 years and OR_10_ is the effect of educational attainment ≤12 in those with no DNA methylation age acceleration

^*^ *p* < 0.05, ^**^ *p* < 0.01, ^***^ *p* < 0.001

**Supplemental Table 5**: Four-way effect decomposition results for high education and no DNA methylation age acceleration based on the DunedinPoAM38 method on Power’s dementia algorithm classification in the Health and Retirement Study, 2016 wave

|  | **DunedinPoAM38** | | | |
| --- | --- | --- | --- | --- |
|  | **non-Hispanic White** | | | |
|  | **Excess Effect on OR scale** | **95%CI** | **% Attributable** | **95%CI** |
| **Controlled Direct Effect** | 0.4 | [-0.4,1.2] | 40.4% | [-13.1,94.0] |
| **Interaction Reference** | 0.4 | [0.0,0.9] | 43.3% | [-0.4,87.1] |
| **Interaction Mediation** | 0.1 | [0.0,0.3] | 13.2% | [-0.1,26.6] |
| **Pure Indirect Effect** | 0.0 | [-0.1,0.1] | 3% | [-5.0,11.0] |
| **Total** | 1.0 | [0.1,2.0] |  |  |
| **Portion Mediated** |  |  | 16.2% | [4.6,27.8] |
| **Portion due to Interaction** |  |  | 56.6% | [-0.6,113.7] |
|  | **non-Hispanic Black** | | | |
|  | **Excess Effect on OR scale** | **95%CI** | **% Attributable** | **95%CI** |
| **Controlled Direct Effect** | 0.4 | [-0.4,1.2] | 31.9% | [-17.3,81.2] |
| **Interaction Reference** | 0.7 | [0.0,1.5] | 57.1% | [12,102.2] |
| **Interaction Mediation** | 0.1 | [0.0,0.2] | 8.9% | [1.9,16] |
| **Pure Indirect Effect** | 0.0 | [-0.1,0.1] | 2% | [-3.6,7.7] |
| **Total** | 1.3 | [0.2,2.4] |  |  |
| **Portion Mediated** |  |  | 11% | [4.8,17.1] |
| **Portion due to Interaction** |  |  | 66% | [13.9,118.2] |
|  | **Hispanic** | | | |
|  | **Excess Effect on OR scale** | **95%CI** | **% Attributable** | **95%CI** |
| **Controlled Direct Effect** | 0.4 | [-0.4,1.2] | 37.8% | [-14.7,90.2] |
| **Interaction Reference** | 0.5 | [0.0,1.0] | 47% | [2.6,91.3] |
| **Interaction Mediation** | 0.1 | [0.0,0.3] | 12.5% | [0.7,24.2] |
| **Pure Indirect Effect** | 0.0 | [-0.1,0.1] | 2.8% | [-4.8,10.4] |
| **Total** | 1.1 | [0.1,2.1] |  |  |
| **Portion Mediated** |  |  | 15.3% | [5.2,25.4] |
| **Portion due to Interaction** |  |  | 59.4% | [3.3,115.5] |

Models are adjusted (self-reported race/ethnicity as noted, male, of average age, average percent of granulocytes and monocytes) and weighted to be representative of the United States population.
